# Geographical Variation in COVID-19 Cases, Prevalence, Recovery and Fatality Rate by Phase of National Lockdown in India, March 14-May 29, 2020

**DOI:** 10.1101/2020.06.04.20122028

**Authors:** Ankita Srivastava, Vandana Tamrakar, Moradhvaj, Saddaf Naaz Akhtar, Krishna Kumar, Tek Chand Saini, C Nagendra, Nandita Saikia

## Abstract

**Background:** Since the COVID-19 pandemic hit Indian states at varying speed, it is crucial to investigate the geographical pattern in COVID-19. We analyzed the geographical pattern of COVID-19 prevalence and mortality by the phase of national lockdown in India.

**Method:** Using publicly available compiled data on COVID-19, we estimated the trends in new cases, period-prevalence rate (PPR), case recovery rate (CRR), and case fatality ratio (CFR) at national, state and district level.

**Findings:** The age and sex are missing for more than 60 percent of the COVID-19 patients. There is an exponential increase in COVID-19 cases both at national and sub-national levels. The COVID-19 infected has jumped about 235 times (from 567 cases in the pre-lockdown period to 1,33,669 in the fourth lockdown); the average daily new cases have increased from 57 in the first lockdown to 6,482 in the fourth lockdown; the average daily recovered persons from 4 to 3,819; the average daily death from 1 to 163. From first to the third lockdown, PPR (0.04 to 5.94), CRR (7.05 to 30.35) and CFR (1.76 to 1.89) have consistently escalated. At state-level, the maximum number of COVID-19 cases is found in the states of Maharashtra, Tamil Nadu, Delhi, and Gujarat contributing 66.75 percent of total cases. Whereas no cases found in some states, Kerela is the only state flattening the COVID-19 curve. The PPR is found to be highest in Delhi, followed by Maharastra. The highest recovery rate is observed in Kerala, till second lockdown; and in Andhra Pradesh in third lockdown. The highest case fatality ratio in the fourth lockdown is observed in Gujarat and Telangana. A few districts viz. like Mumbai (96.7); Chennai (63.66) and Ahmedabad (62.04) have the highest infection rate per 100 thousand population. Spatial analysis shows that clusters in Konkan coast especially in Maharashtra (Palghar, Mumbai, Thane and Pune); southern part from Tamil Nadu (Chennai, Chengalpattu and Thiruvallur), and the northern part of Jammu & Kashmir (Anantnag, Kulgam) are hot-spots for COVID-19 infection while central, northern and north-eastern regions of India are the cold-spots.

**Conclusion:** India has been experiencing a rapid increase of COVID-19 cases since the second lockdown phase. There is huge geographical variation in COVID-19 pandemic with a concentration in some major cities and states while disaggregated data at local levels allows understanding geographical disparity of the pandemic, the lack of age-sex information of the COVID-19 patients forbids to investigate the individual pattern of COVID-19 burden.

**Major highlights of the study:**

i. The new cases of COVID-19 have increased exponentially since the second lockdown phase in India. There is consistent improvement in the recovery rate (CRR is 7.1 percent in pre-lockdown to 44.0 percent in fourth lockdown period) with a low level of CFR (1.87 percent as of May 29^st^ 2020).
ii. At the state level, the most vulnerable states for the COVID-19 crisis are the state of Maharashtra, Tamil Nadu, Delhi, and Gujarat contributing 66.75 percent of total cases.
iii. The PPR is found to be highest in Delhi, followed by Maharastra. While the highest recovery rate is observed in Kerala, the highest case fatality ratio in the fourth lockdown is observed in Gujarat and Telangana. The top 10 hotspot districts in India account for 58.3 percent of the new cases. Among them, Mumbai has the highest infection rate of 96.77 per 100 thousand, followed by Chennai with 63.66 per 100 thousand, and Ahmedabad with 62.04 per 100 thousand.
iv. The information on age and sex are missing for more than 60 percent of the patients.

## 1. Introduction

The novel Coronavirus 19 (COVID-19), caused by severe acute respiratory syndrome, has been spreading rapidly across the world since it broke out in Wuhan city, China in December 2019. COVID-19 outbreak is a Global Public Health Emergency and declared as a global pandemic on March 11, 2020 (Sohrabia et al., 2020; WHO, 2020). On May 29, 2020, this disease has been transmitted to over 213 countries and territories affecting more than 6.24 million people and claiming 0.37 million lives worldwide (Worldometers, 2020). Out of all these 213 countries, areas, or territories, the United States now has the largest outbreak of COVID-19 with 1.8 million (Worldometers, 2020).

In India, the first case of COVID-19 was detected on January 30, 2020, in Kerala (MoHFW, 2020). With the experience of the overwhelming burden of COVID-19 in European countries, the Government of India (GOI) was quick to respond to COVID-19 by a number of preventive measures such as tracking individuals with international and national travel history, quarantining COVID-19 patients, canceling visas and international flights, involving media in raising mass awareness on preventive measures and finally a complete nationwide lockdown from March 24, 2020 to May 17, 2020, by a phase-wise declaration. Later, GOI also introduced a smartphone-based application, known as “Aarogya Setu” to trace and detect COVID-19 affected cases. To reduce the peace of contamination, administration across the country sealed and sanitized the areas or housing societies with COVID-19 patients.

Despite all these measures, the number of COVID-19 cases has been rapidly expanding but with a huge geographical disparity with zero active cases in the northeastern states to 35,058 cases in Maharashtra (on May 19, 2020) (COVID19 INDIA, 2020). Examining the geographical variation in COVID-19 is crucial for understanding the national and local burden of this pandemic. Such a study can provide input for intervention and policy and indicate the future trajectory of COVID-19. The aim of this paper is to analyze the trajectory of COVID-19 cases, period prevalence rate, recovery, and fatality rate by phase-wise during national lockdown at national and sub-national levels.

There are a few non-peer reviewed studies in India addressing COVID-19. Using 310 deaths and 2,519 confirmed cases, a study examined the effect of COVID-19 on longevity in India and inferred that India may lower the life expectancy by 0.3, 0.7 and 1.3 years under various assumptions (Mohanty & Sahoo, 2020). The highest observed CFR among most affected states is in Maharashtra, whereas the recovery rate has reached 72 percent in Kerala, 56 percent in Haryana and 32 percent in Tamil Nadu (Dhillon et al., 2020). A study on spatial and demographic characteristics of COVID-19 positive cases in India shows that majority of the COVID-19 cases are concentrated in Maharashtra, Tamil Nadu, Gujarat, and New Delhi (Ram, Kumar, & Kaur, 2020). The 20–59 aged are the adversely affected groups and two– thirds of the positive cases in India were males(Ram et al., 2020). If the basic reproduction rate during the second lockdown continues, only a maximum of 1.5 percent population will be infected by the middle of August 2020 and the prevalence is likely to reach zero by November 2020 (Kumar, Meitei, & Singh, 2020). A study has used a mathematical model of the spread of COVID-19 in India for both age and social contact structure, where the mortality due to COVID-19 infection showed significant gaps across the age groups, except among elderly where mortality rates rise (R. Singh & Adhikari, 2020) The preventive measures may have averted around 63,334 confirmed cases and 3,845 deaths till April 23, 2020 (Dwivedi, Rai, Shukla, Dey, & Ram, 2020).

The aim of our study is of two-folds: first, we analyzed the national and sub-national trajectory of COVID-19 cases, period-prevalence rate, fatality, and recovery rates by lockdown phases and Secondly, we analyzed the geographical clustering of COVID-19 infection rates in India.

## 3. Data on COVID-19 infection and deaths in India

We used publicly available data from https://www.covid19india.org/ (COVID19INDIA, 2020). It is an Application Programming Interface (API) for daily monitoring of the COVID-19 cases at national, state, and district levels. Data compiled in this web portal is based on state bulletins and official handles to update the case numbers. The details of the data are available on the website. This portal data matches with the data provided by the Ministry of Health and Family Welfare, Government of India (https://www.mohfw.gov.in/) (MoHFW, 2020).

### Missing information on patient’s age and sex

We found that age information is missing for 57 percent of the patients, whereas sex information is missing for 60 percent of the total confirmed cases. Out of the total deceased, about 70 percent do not have information on age and sex. Out of recovered, only 2 percent have age information and about 3 percent have information on sex (See Table 1).

**Table 1.** Age and sex missing case by current status of COVID19 patients India, Jan 30, 2020 to May 29 2020.

|  |  |  |  |
| --- | --- | --- | --- |
| **Current Status** | **Age missing** | **Sex Missing** | **Total Cases** |
| Deceased | 1317 (77.4%) | 1309 (76.9%) | 1702 |
| Hospitalized | 34,665(56.3%) | 32,188 (52.2%) | 61,592 |
| Recovered | 5,651 (97.4%) | 5,567 (95.9%) | 5,801 |
| **Total** | **41,865 (60.4%)** | **39,296 (56.6%)** | **69,329** |

Note: Calculation based on raw data files from https://www.covid19india.org/ website.

Due to such a high level of missing information on age-sex data, we did not carry out our analysis by age-sex disaggregation. About 2.7% of the patients were not assigned to any state, which is excluded from the analysis.

## 4. Methods

### Measures

We analyzed the trends in cases and average daily cases during lockdown periods at national and state levels. We calculated the period prevalence rate (PPR), the case recovery rate (CRR), and the case fatality ratio (CFR), using the following methods:

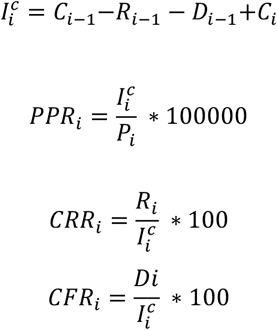

where; Ii^c^ is total COVID-19 infected people in the i^th^ lockdown; Ci is the number of infected cases in the i^th^ lockdown *i*; Ri-1 is the number of recovered in the i-1^th^ lockdown; ṙ_і−1_ is the number of deaths i-1^th^ lockdown; CFRi is case fatality ratio in i^th^ lockdown period; CRRi is case recovery rate i^th^ lockdown; PPRi is the period prevalence rate in the i^th^ lockdown and Pi is the total population of a specific area in i^th^ lockdown.

Since district level data also provides the number of active cases (cases which has not been recovered/deceased and currently infected) of COVID-19, we computed infection rate as

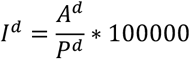

where, I^d^ is the infection rate for particular district d; A^d^ is the number of active cases in district d and P^d^ is the total population of district d.

**Fig 1:**
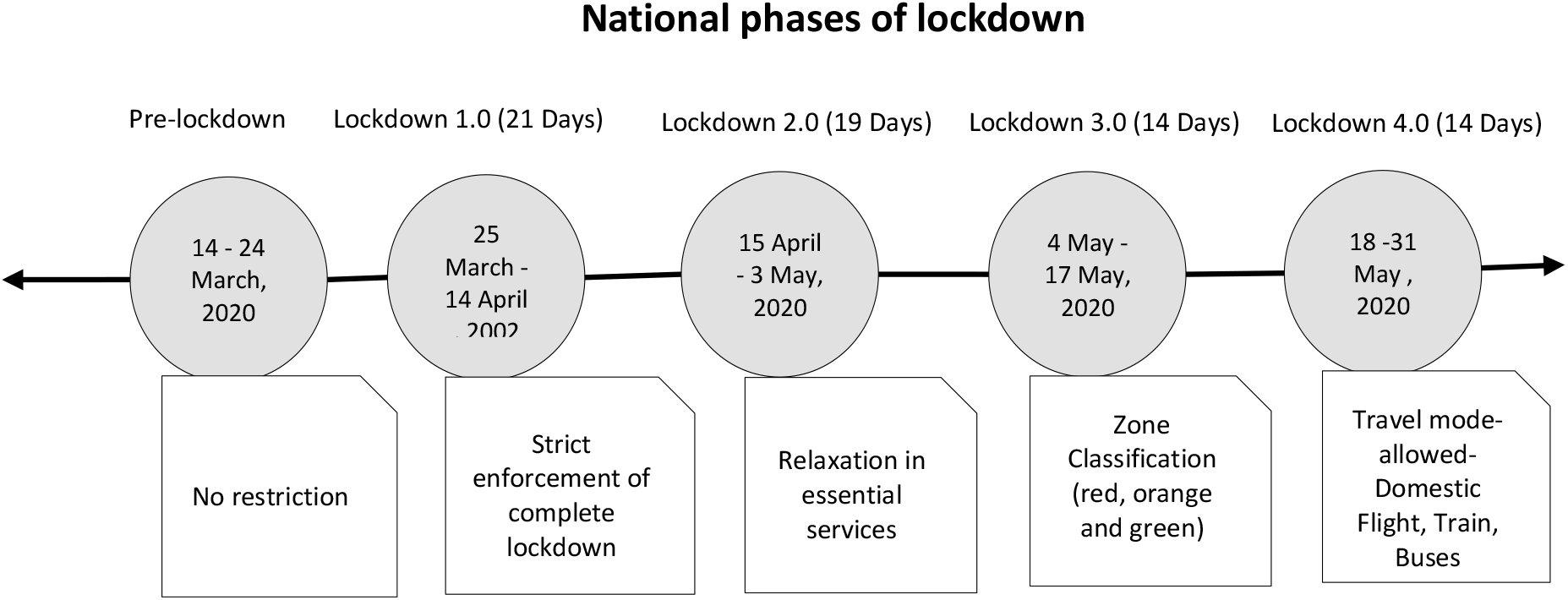
Graphical presentation of various phases of national lockdown in India 14 March, 2020–31 May, 2020.

We carried out the analysis by the phase of the national lockdown that GOI imposed. Figure 1 presents the timeline of lockdown phases: pre-lockdown as March 14, 2020-March 24, 2020 (ten days); first lockdown as March 25, 2020-April 14, 2020 (21 days); second lockdown as April 15, 2020-May 3, 2020 (19 days), third lockdown as May 4, 2020-May 17, 2020 (14 days) and, fourth lockdown as May 18, 2020 – May 31, 2020 (14 days)*, we analysed upto 29^th^ May 2020.

### Geo-spatial analysis

We used QGIS software to generate descriptive maps and later exported shapefiles to GeoDa software to perform spatial analysis. We used first-order Queen’s contiguity matrix as the weight for conducting spatial analysis since our main aim is to understand spatial interdependence between infection rate and the neighboring natural regions. We estimated Moran’I and univariate Local Indicators of Spatial Association (LISA). Moran’s I is the Pearson coefficient measure of spatial autocorrelation, which measures the degree to which data points are similar or dissimilar to their spatial neighbors (Moran, 1950). The LISA cluster map yields four types of geographical clustering of the interest variable (Weinreb, Gerland, & Fleming, 2008). Here, “high-high” means that regions with above-average infection rates also share boundaries with neighboring regions that have above average values of infection rate. On the other hand, “high-low” means that regions with above-average infection rate are surrounded by regions with below-average values. The “high-high” are also referred to as *hot spots*, whereas the “low-low” are referred to as *cold spots*. Our analysis was done in R, GeoDa, and QGIS software.

## 5. Results

### 5.1 National level pattern

India has experienced exponential growth of new, recovered, and deceased cases since the second lockdown phase (Fig 2). The average daily new cases have increased from 57 in the first lockdown to 6,482 in the fourth lockdown; the average number of daily recovered persons from 4 to 3,819; the average number of daily deaths from 1 to 163. The average daily total infected people raised from 57 in the first lockdown to 11,139 in the fourth lockdown. There is rise in new cases (567 to 77,790); total infected persons (567 to 1,33,669); recovered (40 to 45,832) and deaths from 10 to 1,955 (See Fig 2).

**Fig 2:**
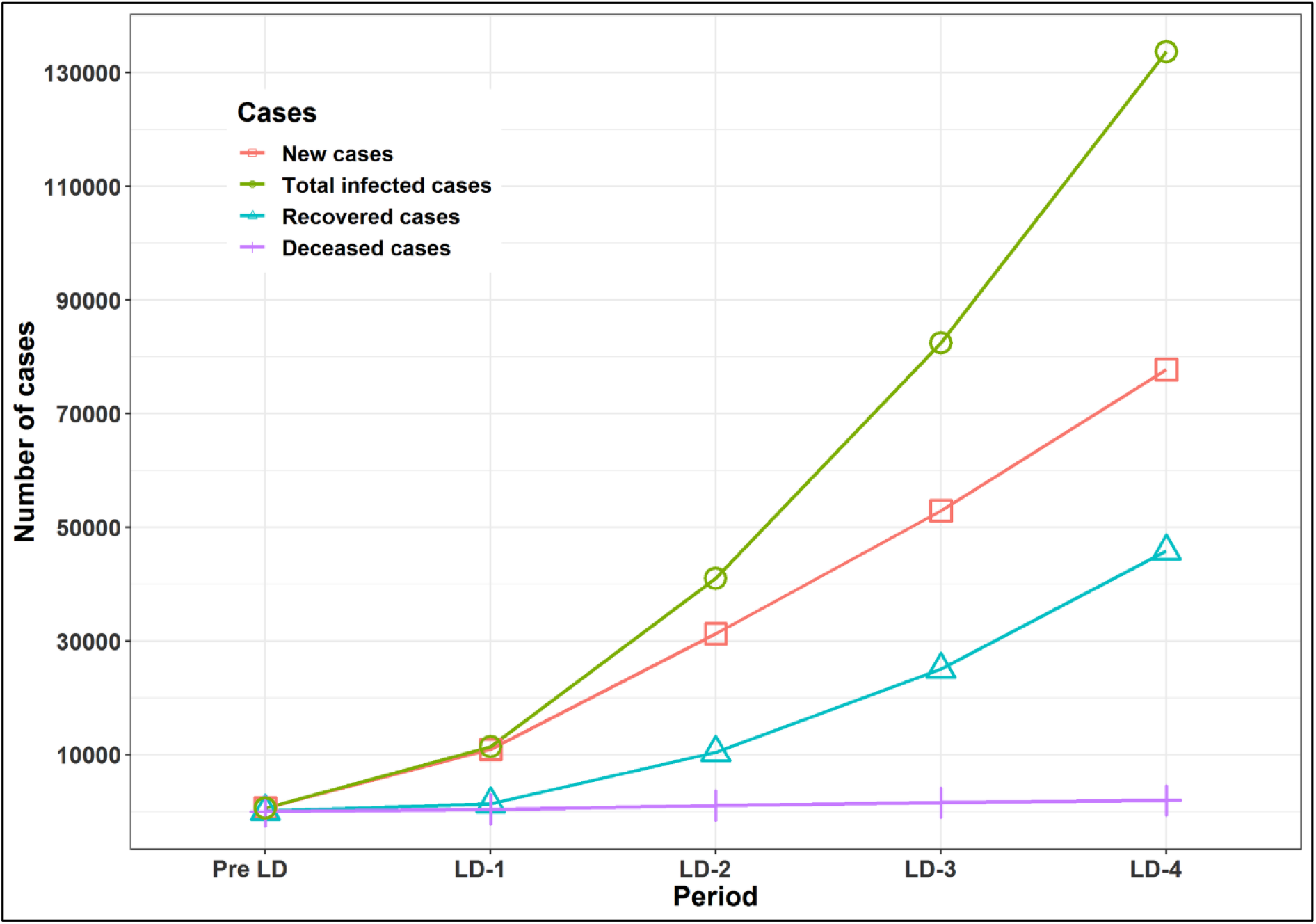
New, total infected, recovered and deceased cases of COVID-19 by lockdown phases in India, March 14-May 29, 2020.

Fig 3 shows PPR per 100 thousand, CRR per 100, and CFR per 100 by lockdown phases in India. It is evident that at national level PPR (0.04 to 9.64), CRR (7.05 to 34.29) has consistently increased over these periods whereas CFR (1.76 to 1.46) has declined marginally (Fig 3).

**Fig 3:**
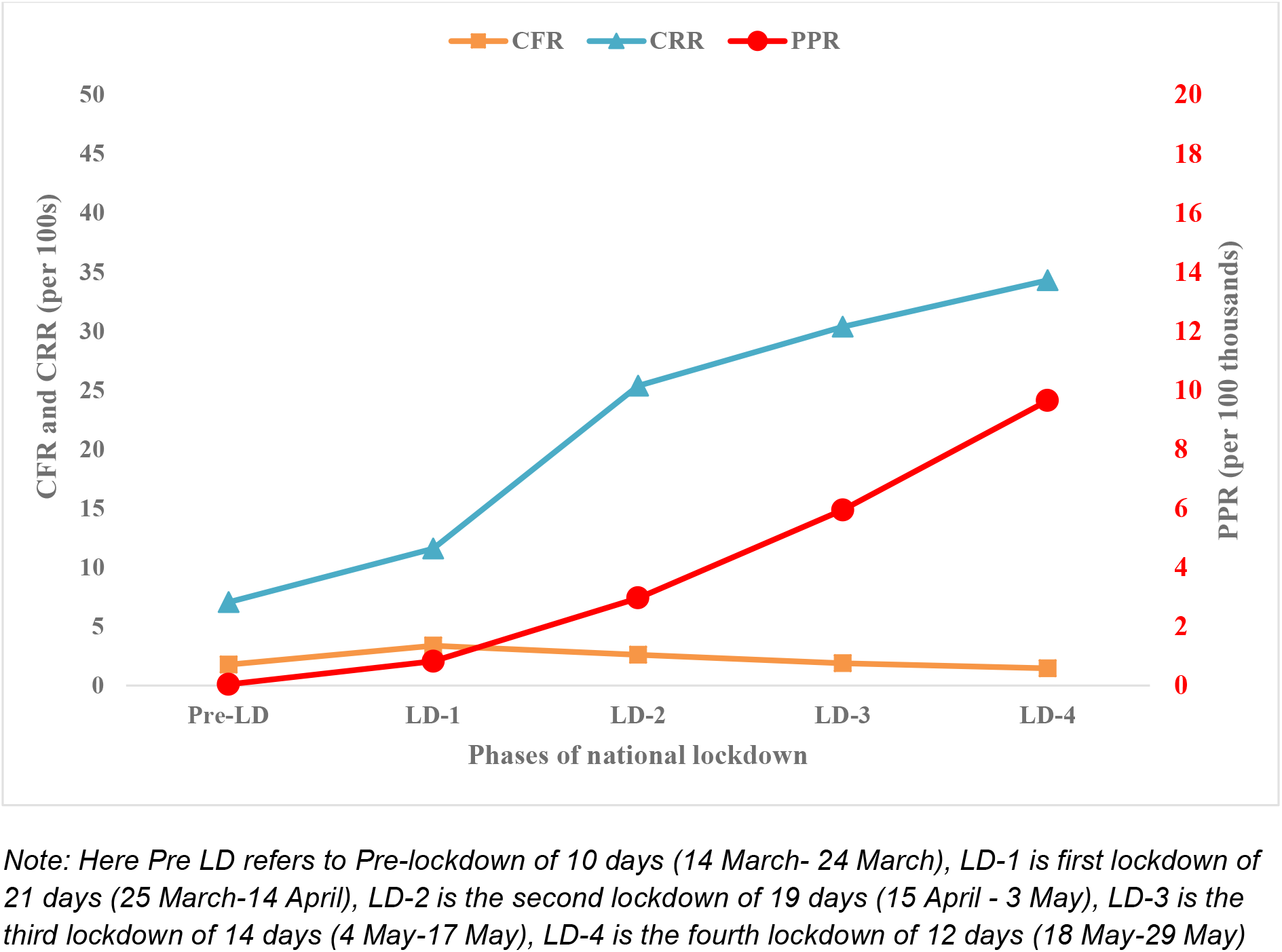
Period prevalence rate (PPR per 1,00,000), Case Recovery Rate (CRR per 100) and Case fatality rate (CFR per 100), in COVID-19 by lockdown phases in India, March 14-May 29, 2020.

### 5.2 State level pattern

Fig 4 and S3 Table present the state-level pattern of new, total infected, recovered, and deceased cases of COVID-19. The maximum new cases were found in Maharashtra, whereas no case was found in a few smaller states/UT_s_ (Daman & Diu; Lakshadweep; Mizoram and Andaman & Nicobar Islands) until the end of the fourth lockdown period. The average daily and the total number of new cases has increased consistently nearly in all states over the lockdown period. Maharashtra (from 11 to 2,431 persons per day and total new cases from 107 to 29,175); Tamil Nadu (average 2 to 751 persons per day and total new cases from 18 to 9,022); Delhi (average 3 to 635 persons per day and total new cases from 30 to 7,631); Gujarat (average 3 to 380 persons per day and the total new cases from 34 to 4,564); Rajasthan (average 3 to 263 persons per day and total new cases 32 to 3163). However, Kerala is the only state experiencing a decline in new cases per day till the third lockdown (average 11 to 8 persons per day and the total new cases 109 to 102), however, Kerala experience a spike in the fourth lockdown (average 7 to 45 new cases per day and total new cases 102 to 549). Among northeastern states, Assam (0 to 79 persons per day and total new cases 0 to 957) has the highest number.

**Fig 4:**
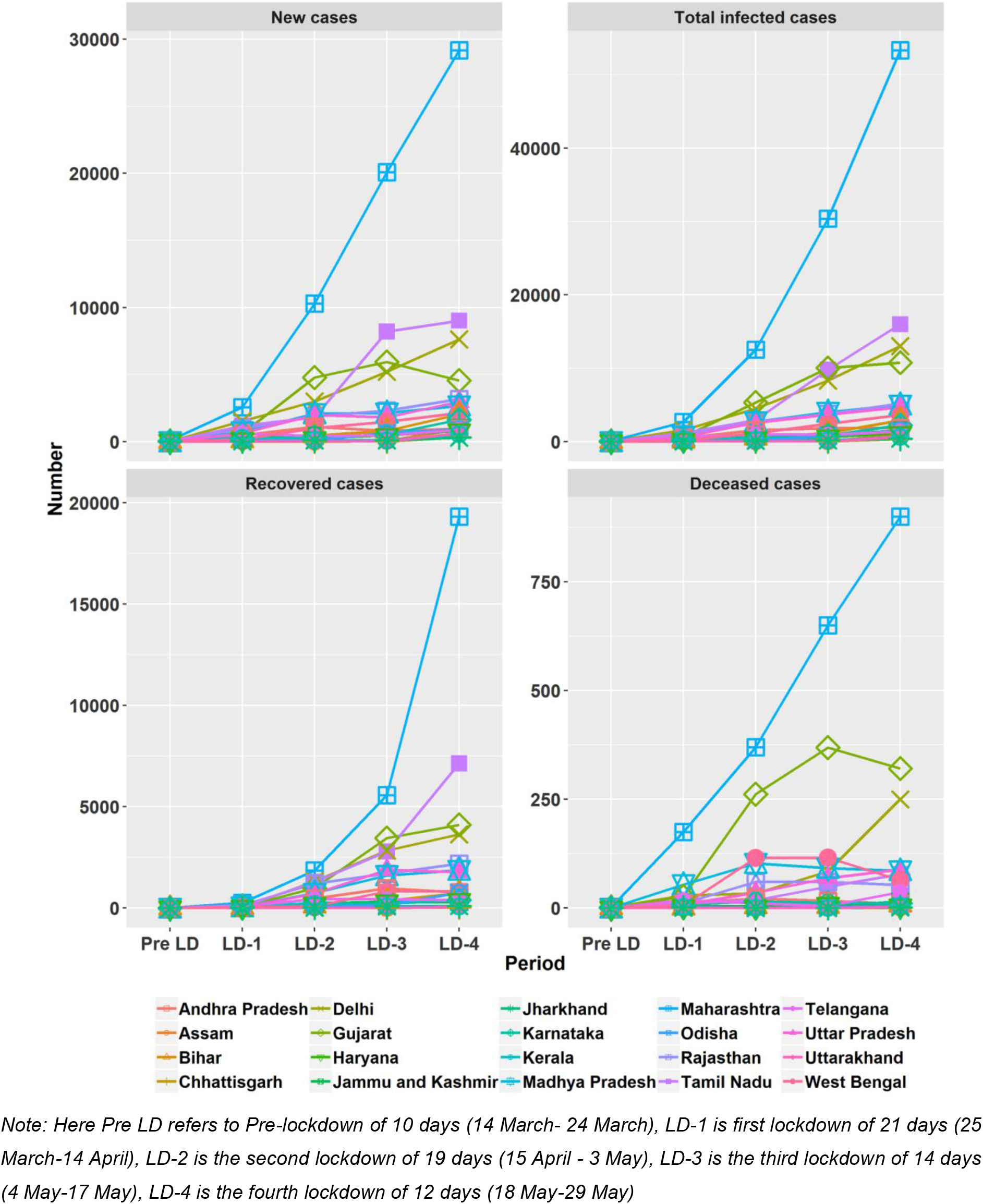
State-wise new, total infected, recovered and deceased COVID-19 cases by lockdown phases in India, March 14-May 29, 2020.

Interestingly, overall 66.75 percent of COVID-19 total infected persons belong to the four states, viz., Maharashtra, Tamilnadu, Delhi, and Gujarat. Also, the average daily prevalence of COVID-19 is the highest in the six major states and has increased consistently viz. Maharashtra (11 to 4,445 persons per day; total infected persons 107 to 53,343); Tamil Nadu (2 to 1,332 persons per day; total infected persons 18 to 15,995); Delhi (3 to 1086 persons per day; total infected persons 30 to 13,036); Gujarat(3 to 899 persons per day; total infected persons 34 to 10,786); Rajasthan (3 to 431 per day; total infected persons 32 to 5,179) and Madhya Pradesh (1 to 416 per day; total infected persons 7 to 4,993), though Kerala experienced the least increment in average prevalence of COVID-19 (11 to 54 per day; 109 to 650 total persons) at the end of the fourth lockdown(see Fig 4).

State-wise trend analysis of recovered cases also varied during lockdowns. The highest recovered cases belong to Maharashtra and the lowest to Meghalaya. Besides, the highest daily average and number of recovered cases belong to Maharashtra (0 to 1609 average daily cases; recovered from 0 to 19,309); Tamil Nadu (0 to 595 average daily cases; recovered from 1 to 7,141); Gujarat (0 to 342 average daily; recovered from 0 to 4,112); Delhi (1 to 236 average daily cases; recovered from 6 to 2,840); Rajasthan (0 to 182 average daily cases; recovered from 3 to 2,189). Kerala experienced a decline in average daily recovered cases due to smaller number of infected cases in the fourth lockdown period.

The state variation in deaths has been seen from one to another lockdown phase. Maharastra recorded the highest number of deaths irrespective of the lockdown phase. In the first phase of lockdown, the second-highest death cases were found in Madhya Pradesh (54 deaths) followed by Delhi (29 deaths) and Gujarat (27 deaths) whereas in second and third lockdown phases, Gujarat and West Bengal ranked the second position respectively. In the fourth phase, Gujarat (321) and Delhi (250) experienced the highest spike in deaths after Maharashtra.

Maharashtra (average daily deaths from 0 to 75; deaths from 2 to 900) also experienced the highest number of deaths over the lockdown period, followed by Gujarat (average daily deaths from 0 to 27; deaths 1 to 321); Delhi (average daily deaths from 0 to 21; deaths from 1 to 250) and the lowest in Kerala with no deaths in third lockdown to 5 deaths in fourth lockdown.,

However, PPR per 100 thousand population is the highest for Delhi (65.79) and the lowest for Jharkhand (1.08) in the fourth lockdown among the top 20 infected states (Figure 5). The pattern of PPR by states has increased consistently in four states/UTs viz., Delhi (0.15 to 65.79); Maharashtra(0.09 to 43.67); Tamil Nadu (0.02 to 21.13); Gujarat (0.05 to 15.88); Jammu & Kashmir (0.05 to 11.94). Though in Kerala (0.31 to 1.85) PPR has slightly increased compared to other states. (see Fig 5).

**Fig 5.**
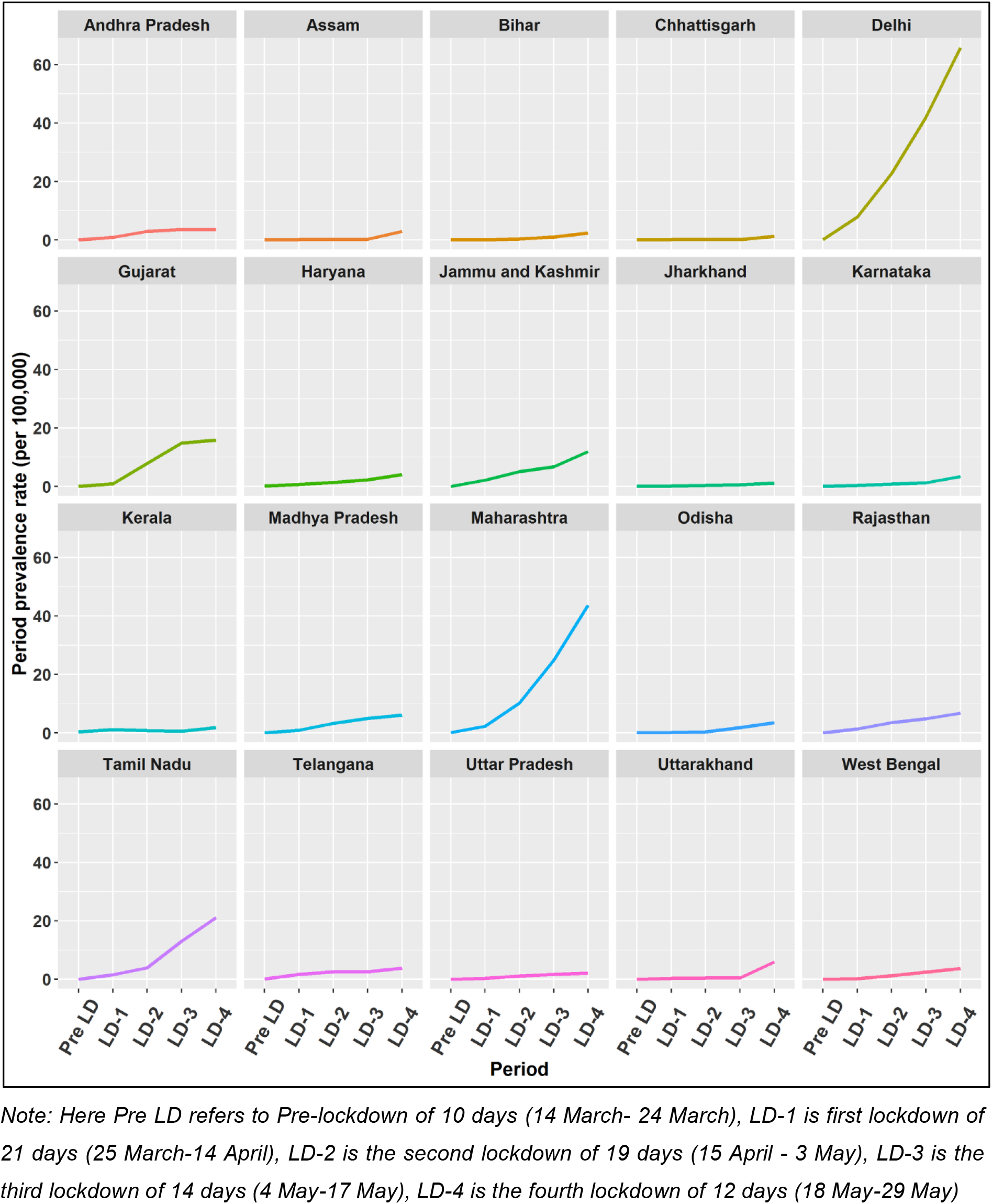
State-wise period prevalence rate (PPR) per 100 thousand by lockdown phases in India, March 14-May 29, 2020.

At statelevel, the CRR (per 100s) varied across states. Among top 20 infected states, Odisha (50.50); Tamil Nadu (44.65); Andhra Pradesh (42.65); Rajasthan (42.27); Gujarat (38.12); Uttar Pradesh (37.55) observed the highest recovery rate and Uttarakhand (7.54) observed the lowest recovery rate in fourth lockdown period (See Fig 6). The pattern of CRR varied from the first to fourth phase of lockdown. While most of the states observed an inverted “U” pattern, some states such as Gujarat, Madhya Pradesh, Maharashtra, Orissa, and Andhra Pradesh experienced increasing recovery rates. On an average Haryana (37.99) showed the highest recovery rate followed by Kerala (36.67) and Rajasthan (31.07) over all lockdowns.

**Fig 6.**
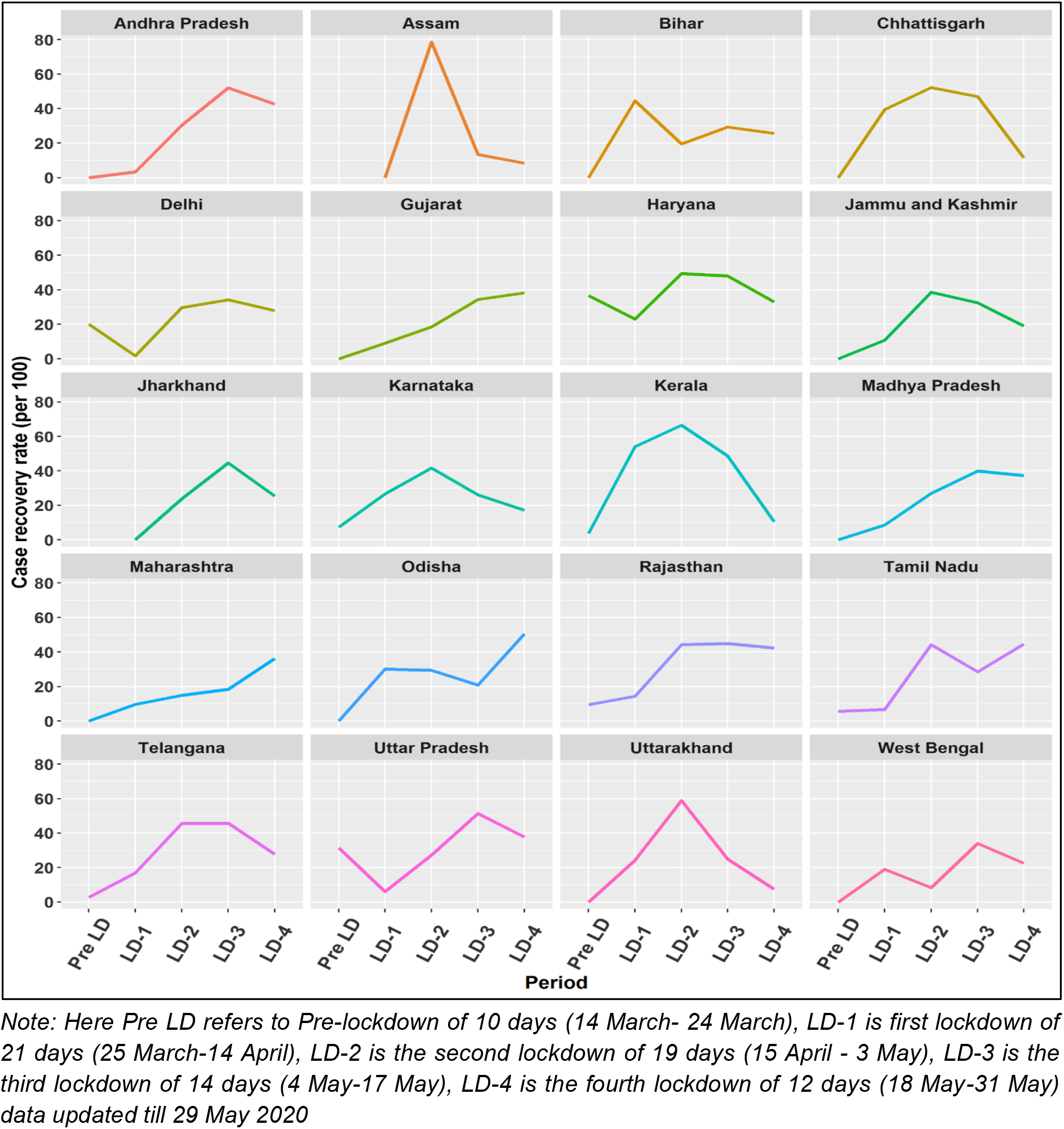
State-wise Case Recovery Rate (CRR) (per 100s) by lockdown phases in India, March 14-May 29, 2020.

Among the states with at least 100 infected cases, CFR is the highest in Gujarat (2.98); Telangana(2.64); Delhi (1.92); Uttar Pradesh (1.89) and the lowest in Assam (0.20) and Bihar (0.24) in the fourth lockdown. Pattern of CFR showed that other larger states have also experienced high fatality over the lockdown periods viz; Telangana (0 to 2.64); Uttar Pradesh (0 to 1.89); Madhya Pradesh (0 to 1.72); Rajasthan (0 to 1.02). However, we found a few states experiencing decline in fatality rate, viz. Bihar(33.3 to 0.24); West Bengal (11.1 to 1.77); Tamil Nadu (5.5 to 0.49); Karnataka (2.4 to 0.49); Delhi (3.3 to 1.92) and, Maharashtra (1.87 to 1.69) (see Fig 7).

**Fig 7.**
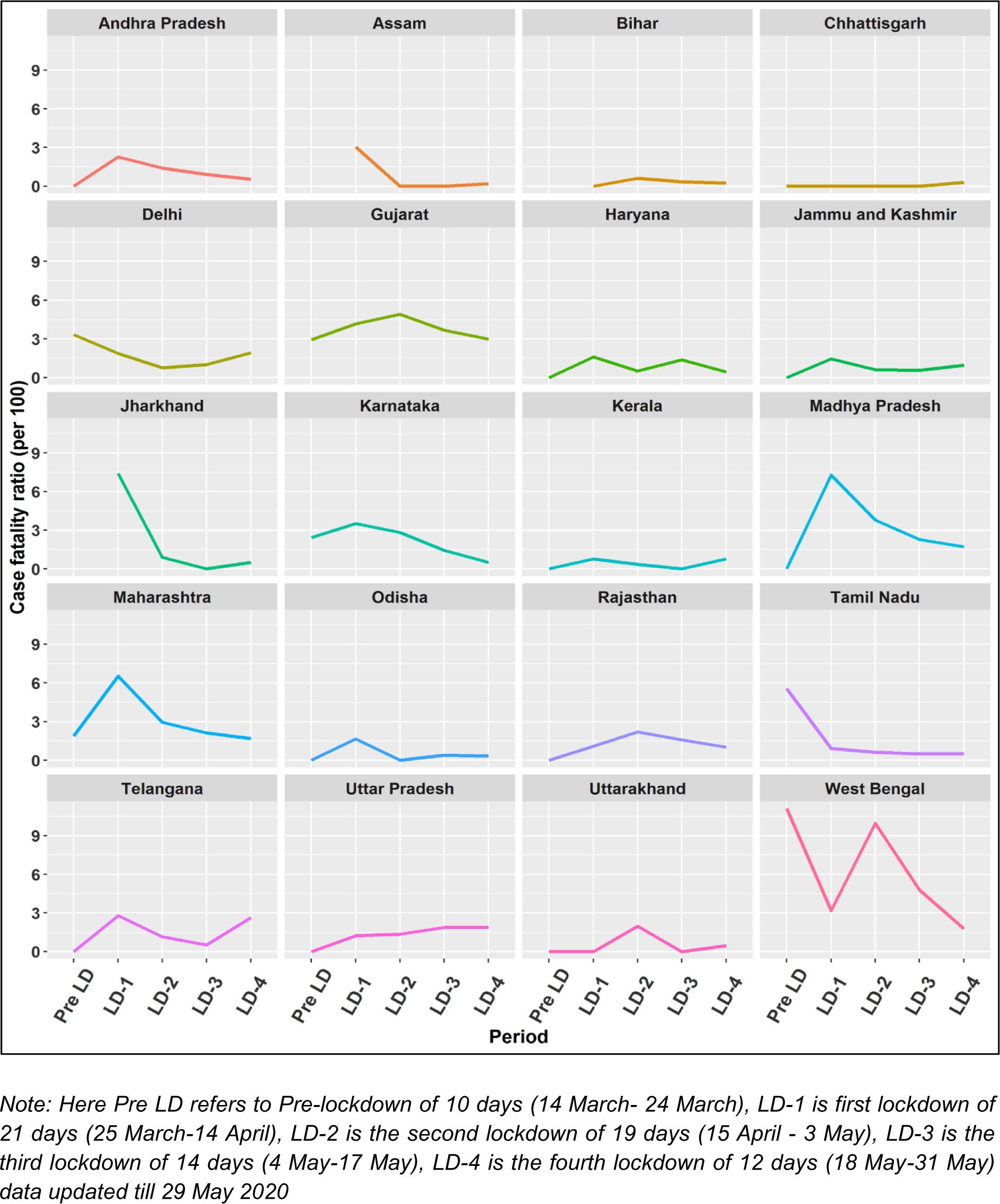
State-wise Case Fatality Ratio (CFR) per 100 by lockdown phases in India,March 14-May 29, 2020.

### 5.3 Spatial variation of COVID-19 positive cases by districts, till May 29, 2020

Fig 8 district-level COVID-19 situation in India. The size of the bubble indicates the number of positive COVID-19 cases at districts on May 29, 2020. The larger the size of the bubble is the higher the positive cases. Our study observes that out of 736 districts, just four districts contributes about 47 percent confirmed cases (Mumbai; Chennai; Ahmedabad and Thane). Out of these districts, about 88 percent (648 districts) have at least one case of COVID-19 and about 25 percent districts (182 districts) with at least 100 patients. The top five affected districts with the highest positive cases till the fourth lockdown are Mumbai, Chennai, Ahmedabad, Thane, and Pune.

**Fig 8.**
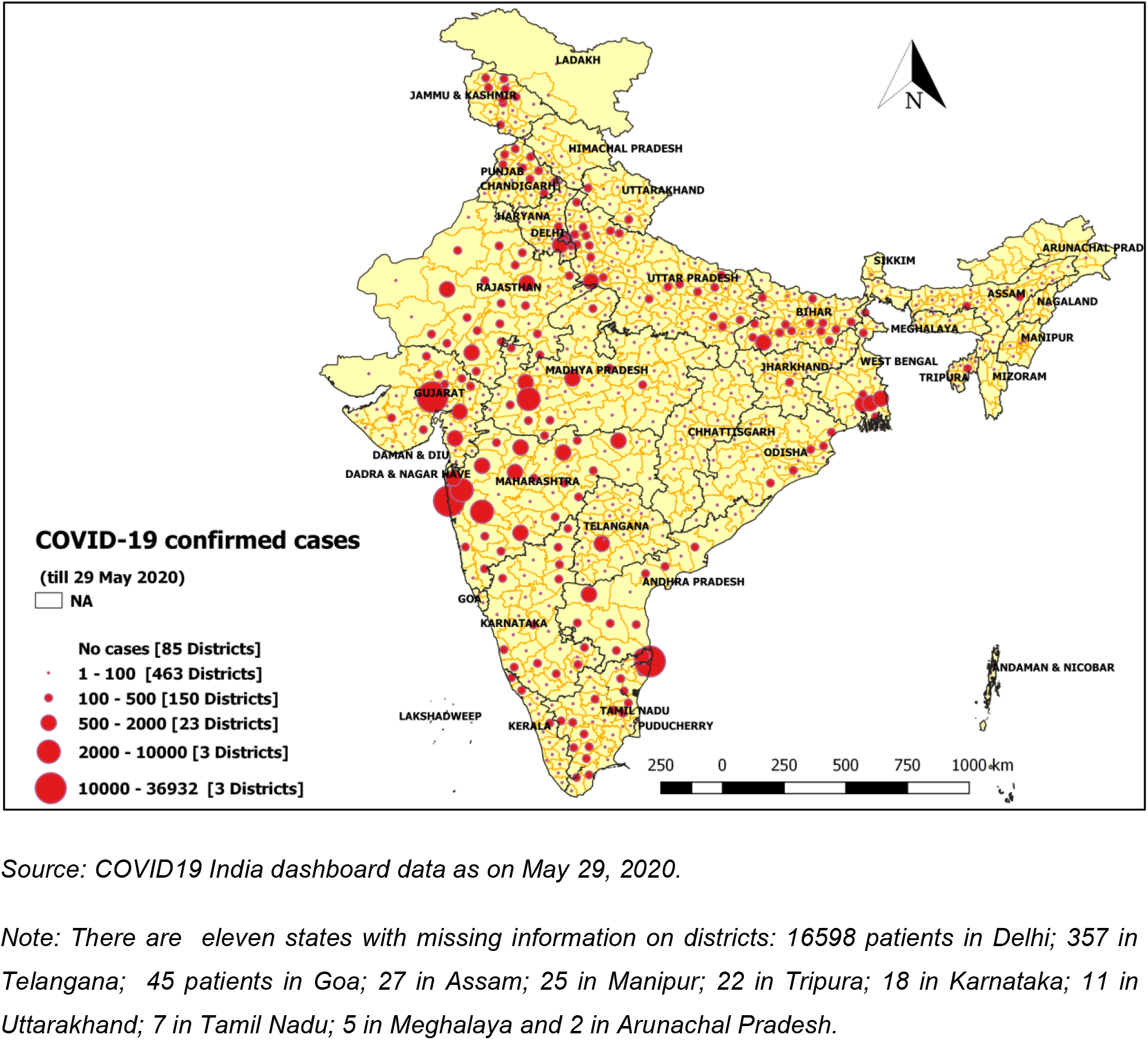
Spatial variation of COVID-19 positive cases by zones/districts on May 29, 2020.

Fig 9 presents the district level infection rate, defined as the number of active cases per 100 thousand population by May 29, 2020. Results show that the top 10 hotspot districts in India account for 30 percent of the cases. Twelve districts have an infection rate above 20 per 100 thousand population. These are Mumbai, Ahmedabad, Chennai, Ariyalur, Palghar, Indore, Aurangabad, Central Delhi, Dhalai, New Delhi, Perambalur, Thane, Pune, and Agra. Among those, Mumbai (96.7 per 100 thousand); Chennai (63.66 per 100 thousand); Ahmedabad (62.04 per 100 thousand); Indore (51.21 per 100 thousand) has the highest infection rate among districts. Incidentally, most of these are big Indian affluent cities, which are also powerful economic zones. The Moran’s I value of 0.104 representing a lower but positive level of spatial clustering in case of the COVID-19 infection rate over neighbouring districts. While clusters in the parts of Konkan coast especially in Maharashtra (Palghar, Mumbai, Thane, and Pune); the southern part from Tamil Nadu (Chennai, Chengalpattu, and Thiruvallur) and the northern part of Jammu & Kashmir (Anantnag, Kulgam) are found as COVID-19 infection hotspots, cold-spots were observed in central, northern and north-eastern regions of India.

**Fig 9:**
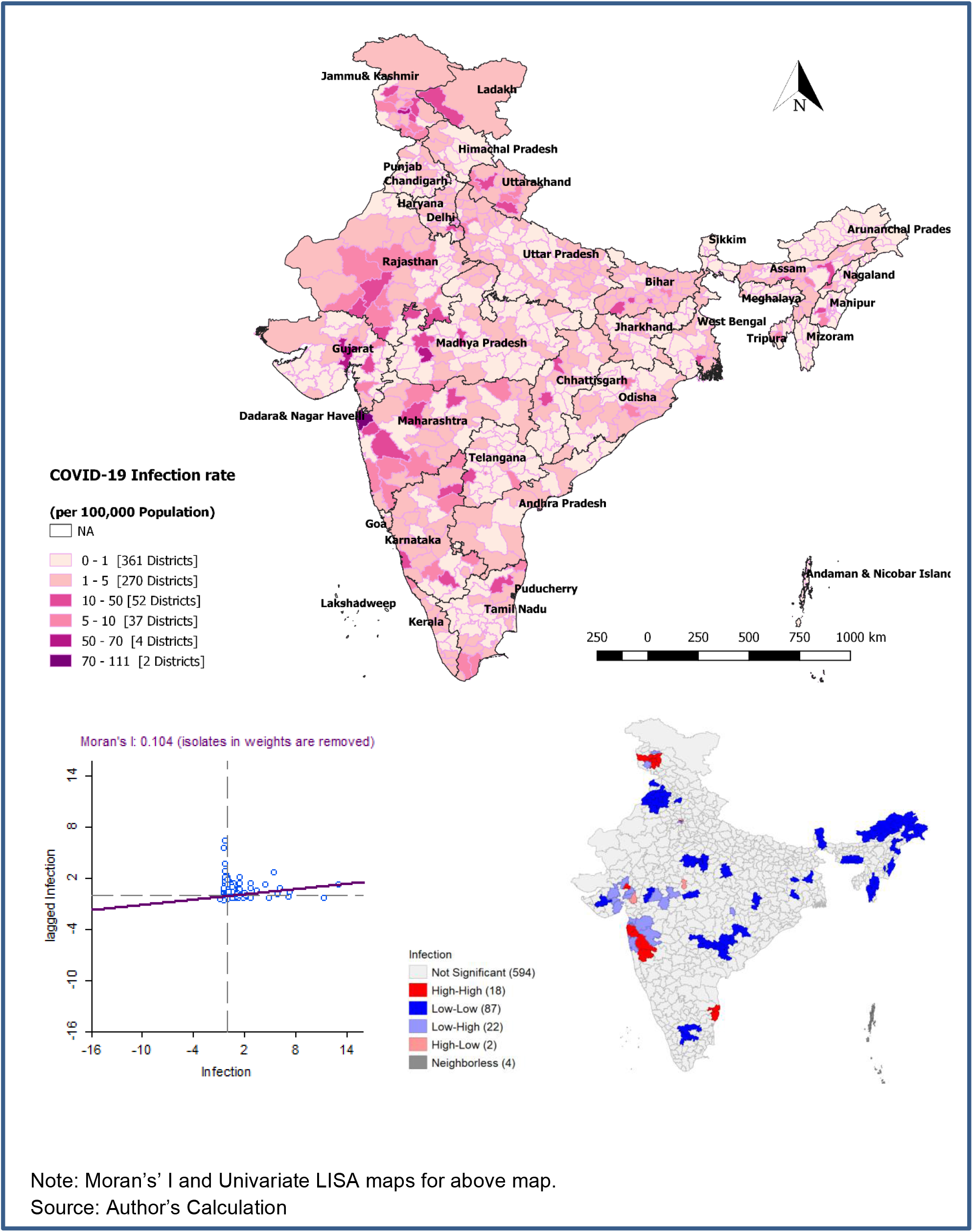
District level spatial pattern of COVID-19 infection rate till fourth lockdown (May 29, 2020).

## 6. Discussion and conclusion

COVID-19 is a global challenge where India stands within the first ten affected countries. The share by India to COVID-19 cases may arise in the near future because of its mammoth population size, high population density, and poor public health facilities. The present study provides the national and sub-national trajectory of COVID-19 cases, period-prevalence rate, fatality rate and recovery rate in India, over a period of four months, from onset to May 29, 2020 (end of the nationwide fourth lockdown).

As of May 29, 2020 (by end of 4^th^ National lockdown), India has reported a total of 1,73,487 cases, 82,627 recovered and 4,977 deaths from COVID-19 in 37 states and union territories. The national daily new cases observed about 97 percent change over lockdown period; daily infected about 130 percent change; daily recovered about 817 percent change and daily deceased about 139 percent change. Our analysis also revealed that at the national level, there is consistent improvement in CRR over the lockdown period. We also found the CFR was minimal by the end of the fourth lockdown phase.

This study also observed that at the national level, the PPR has increased 183 times over four lockdown periods The highest percent change is observed in Bihar (828 times); Tamil Nadu (797 times); Odisha (698 times); Madhya Pradesh (443 times) from first to fourth lockdowns; whereas, the percent change of CRR is the highest in Telangana (15.0 percent change); Tamil Nadu (8 percent change); Rajasthan(5 percent change) and Kerala (2 percent change); from first to third lockdown periods. Most of the states observed a decline in CFR over the lockdown periods. However, three states viz., Maharashtra, West Bengal, and Gujarat have to put more effort as they have high CFR and low recovery rates. Maharashtra also observed the lowest CRR of 17.9 percent on average over lockdown periods.

The number of confirmed cases in India is found to be lesser than other countries where lockdown is either not strict or not implemented (Roser, Ritchie, Ortiz-Ospina, & Hasell, 2020). A study found that lockdown has averted 330 deaths until April 23, 2020 (Dwivedi et al., 2020). Thus, lockdown may have played an important role in decelerating the speed. Moreover, it has also given the Government time to prepare for a possible surge in cases, when the pandemic is predicted to peak in the coming weeks. However, PPR rising at an escalated pace (0.4 to 7.5 percent), more importantly in the last two lockdown periods might be due to two possible reasons; firstly, the exercise of testing samples has been intensified by government authorities, which consequently results new cases; secondly, the nation has witnessed huge flux of migrants returning to home states especially from megacities during third and fourth lockdown periods. Moreover, the Home Secretary of India has made some relaxation for the inter-state movement mainly for distressed migrant workers, stranded tourists, pilgrims, and students (GOI, 2020).

The present study observed huge state-level variations among COVID-19 new, total infected, recovered, and deceased cases. Six states (Maharashtra 35.9 percent; Tamil Nadu 11.7 percent; Delhi 10.0 percent; Gujarat 9.2 percent Rajasthan 4.8 percent and Uttar Pradesh 4.3 percent) together account for 75.8 percent of the national new cases, most of them contribute substantially to nation’s GDP. Among the national recovered cases, 66 percent belongs to just four states, percent belong to Maharashtra (33 percent), Tamil Nadu (14 percent), Gujarat (10.0 percent), and Delhi (9.0 percent). At the state-level the pattern of new, total infected, recovered, and deceased cases per day have also increased over the lockdown periods. About 77 percent of COVID-19 deaths belong to four states viz., Maharashtra (42.1 percent), Gujarat (19.7 percent), Delhi (8.0), and Madhya Pradesh (6.7 percent). The percent change of daily new cases and daily total infected cases is the highest in Bihar, Tamil Nadu, Odisha, and Madhya Pradesh. The daily-recovered cases were the highest among Tamil Nadu, Rajasthan, Delhi, and Telangana. The daily deceased cases were the highest in Maharashtra, Gujarat, and Delhi) from the first to the fourth lockdown periods. However, Kerala is the only state experiencing a reverse trend in the daily new cases till third locklockdown but observed 3 times spike in the fourth lockdown.

Kerala comes out to be the best example to fight this contagious disease. It has flattened the curve of this outbreak and observed a consistent decline in CFR (0.7 to 0) till third lockdown periods and reached 48.7 percent recovery rate since the first lockdown period though in fourth lockdown it observed decline in the recovery rate and increase in the fatality rate. Early detection, broad social support (Masih, 2020), together with strong public health system (Kurian, 2020; Vibhute & Chattopadhyay, 2020) helped to contain the virus in Kerala till the third lockdown. Kerala’s previous experience of the Nipah virus in 2018 to use extensive testing, contact tracing, and community mobilization to contain the virus and maintain a very low mortality rate might have also helped. It has also set up thousands of temporary shelters for migrant workers (Lancet, 2020). Another possible reason for lesser infection in Kerala may be lower population density in the cities of Kerala than the cities from the highly affected states. However, the reason for the spike of cases in the fourth lockdown in Kerala might be due to the reemergence of infection due to migration in the state (Hussain, 2020).

The study further finds that the COVID-19 cases are spatially concentrated in the selected districts of India. We observed that out of 736 districts, 648 districts have at least one case of COVID-19 and about 182 districts have at least 100 COVID-19 patients. The pattern of districts revealed that the active COVID-19 cases are mostly concentrated in districts with large population sizes and mostly urbanized. About 30 percent of cases belong to 10 hotspot districts belonging to Maharashtra, Gujarat, Delhi, Madhya Pradesh and Tamil Nadu. Mumbai (96.70 per 100 hundred population) observes the highest infection rate followed by Chennai (63.66 per 100 thousand population) and, Ahmedabad (62.04 per 100 thousand population). The smaller districts such as Ariyalur, Dhalai, Leh, Kurnool, Perambalur also observed a significantly high infection rate of COVID-19. The risk factors of high infection rates may be due to demographic, socio-economic or contextual factors such as age structure, population density, migration rate living conditions, access to health care etc.

The age-sex pattern is crucial to understand the pattern of the pandemic spread and recognize the intensity of public health intervention measures required for disease control. It is missing from the data, about 77 percent for age and about 72 percent from sex. We also observed state-level age-sex missing data.

It is worthwhile to mention that there is a possibility of under-reporting in infected and deceased cases due to lack of diagnosis. Media has highlighted under-reporting of COVID-19 Cases in Delhi and West Bengal (PTI, 2020; S. S. Singh, 2020). By analyzing the available COVID-19 data, we cannot evaluate the total burden of mortality by COVID-19. There might be a spike of deaths among people suffering from fatal diseases like cancer, hypertension because non-COVID healthcare services have been disrupted. Such an indirect effect of COVID-19 on mortality cannot be evaluated since India does not have any data system giving weekly or monthly death records. Nevertheless, data used in the study is the only reliable statistic to understand the geographical disparity in the COVID-19 pandemic in India.

## Data Availability

We used publicly available data from https://www.covid19india.org/. It is an Application Programming Interface (API) for daily monitoring of the COVID-19 cases at national, state, and district levels.

## Notes

### Competing Interest Statement

The authors have declared no competing interest.

### Funding Statement

No funding was received for this work

### Author Declarations

The data is publicly available

